# Transmission, infectivity, and antibody neutralization of an emerging SARS-CoV-2 variant in California carrying a L452R spike protein mutation

**DOI:** 10.1101/2021.03.07.21252647

**Authors:** Xianding Deng, Miguel A Garcia-Knight, Mir M. Khalid, Venice Servellita, Candace Wang, Mary Kate Morris, Alicia Sotomayor-González, Dustin R Glasner, Kevin R Reyes, Amelia S. Gliwa, Nikitha P. Reddy, Claudia Sanchez San Martin, Scot Federman, Jing Cheng, Joanna Balcerek, Jordan Taylor, Jessica A Streithorst, Steve Miller, G. Renuka Kumar, Bharath Sreekumar, Pei-Yi Chen, Ursula Schulze-Gahmen, Taha Y. Taha, Jennifer Hayashi, Camille R. Simoneau, Sarah McMahon, Peter V. Lidsky, Yinghong Xiao, Peera Hemarajata, Nicole M. Green, Alex Espinosa, Chantha Kath, Monica Haw, John Bell, Jill K. Hacker, Carl Hanson, Debra A. Wadford, Carlos Anaya, Donna Ferguson, Liana F. Lareau, Phillip A. Frankino, Haridha Shivram, Stacia K. Wyman, Melanie Ott, Raul Andino, Charles Y. Chiu

## Abstract

We identified a novel SARS-CoV-2 variant by viral whole-genome sequencing of 2,172 nasal/nasopharyngeal swab samples from 44 counties in California. Named B.1.427/B.1.429 to denote its 2 lineages, the variant emerged around May 2020 and increased from 0% to >50% of sequenced cases from September 1, 2020 to January 29, 2021, exhibiting an 18.6-24% increase in transmissibility relative to wild-type circulating strains. The variant carries 3 mutations in the spike protein, including an L452R substitution. Our analyses revealed 2-fold increased B.1.427/B.1.429 viral shedding in vivo and increased L452R pseudovirus infection of cell cultures and lung organoids, albeit decreased relative to pseudoviruses carrying the N501Y mutation found in the B.1.1.7, B.1.351, and P.1 variants. Antibody neutralization assays showed 4.0 to 6.7-fold and 2.0-fold decreases in neutralizing titers from convalescent patients and vaccine recipients, respectively. The increased prevalence of a more transmissible variant in California associated with decreased antibody neutralization warrants further investigation.

## Introduction

Genetic mutation provides a mechanism for viruses to adapt to a new host and/or evade host immune responses. Although SARS-CoV-2 has a slow evolutionary rate relative to other RNA viruses (∼0.8 x10 ^-3^ substitutions per site per year) (Day et al., 2020), an unabating COVID-19 pandemic with high viral transmission has enabled the virus to acquire significant genetic diversity since its initial detection in Wuhan, China in December 2019 (Zhu et al., 2020), thereby facilitating the emergence of new variants (Fontanet et al., 2021). Among numerous SARS-CoV-2 variants now circulating globally, those harboring a D614G mutation have predominated since June of 2020 (Korber et al., 2020), possibly due to enhanced viral fitness and transmissibility (Hou et al., 2020; Plante et al., 2020; Zhou et al., 2021).

Emerging variants of SARS-CoV-2 that harbor genome mutations that may impact transmission, virulence, and immunity have been designated “variants of concern” (VOCs). Beginning in the fall of 2020, 3 VOCs have emerged globally, each carrying multiple mutations across the genome, including several in the receptor-binding domain (RBD) of the spike protein. The B.1.1.7 variant, originally detected in the United Kingdom (UK) (Chand et al., 2020), has accumulated 17 lineage-defining mutations, including the spike protein N501Y mutation that confers increased transmissibility over other circulating viruses (Leung et al., 2021; Rambaut et al., 2020b; Volz et al., 2020). Preliminary data suggest that B.1.1.7 may also cause more severe illness (Davies et al., 2021). As of early 2021, the B.1.1.7 variant has become the predominant lineage throughout the UK and Europe, with reported cases also rising in the United States (US) (Washington et al., 2021). The other two VOCs, B.1.351 detected in South Africa (Tegally et al., 2020) and P.1 first detected in Brazil (Sabino et al., 2021), carry E484K and K417N/K417T in addition to N501Y mutations. Multiple studies have reported that the E484K mutation in particular may confer resistance to antibody neutralization (Cole et al., 2021; Wang et al., 2021; Wibmer et al., 2021; Wu et al., 2021; Xie et al., 2021), potentially resulting in decreased efficacy of currently available vaccines (Liu et al., 2021; Wise, 2021). This phenotype may have also contributed to widespread reinfection by P.1 in an Amazon community that had presumptively achieved herd immunity (Buss et al., 2021; Sabino et al., 2021).

In January 2021, we and others independently reported the emergence of a novel variant in California carrying an L452R mutation in the RBD of the spike protein (CDPH, 2021; Zhang et al., 2021). Here we used viral whole-genome sequencing of nasal/nasopharyngeal (N/NP) swab samples from multiple counties to characterize the emergence and spread of this L452R-carrying variant in California from September 1, 2020, to January 29, 2021. We also combined epidemiologic, clinical, and in vitro laboratory data to investigate transmissibility and susceptibility to antibody neutralization associated with infection by the variant.

## Results

### Viral genomic surveillance

We sequenced 2,172 viral genomes across 44 California counties from remnant N/NP swab samples testing positive for SARS-CoV-2 (**Supplementary Tables 1 and 2**). The counties with proportionally higher representation in the dataset included Santa Clara County (n=725, 33.4%), Alameda County (n=228, 10.5%), Los Angeles County (n=168, 7.7%) and San Francisco County (n=155, 7.1%) (**Figure 1A**). A novel variant, subsequently named 20C/L452R according to the NextStrain nomenclature system (Bedford et al., 2021) or B.1.427/B.1.429 according to the Pango system (Rambaut et al., 2020a) (henceforth referred to using the Pango designation to distinguish between the B.1.427 and B.1.429 lineages), was identified in 21.1% (459 of 2,172) of the genomes (**Supplementary Table 1**). The frequency of this variant in California increased from 0% at the beginning of September 2020 to >50% of sequenced cases by the end of January 2021. The rise in the proportion of sequenced cases due to the variant was rapid, with an estimated increase in transmission rate of the B.1.427/B.1.429 variant relative to circulating non-B.1.427/B.1.429 lineages of 18.6-24.2% and an approximate doubling time of 18.6 days (**Figure 1B**). Similar epidemic trajectories were observed from multiple counties (**Figure 1C-1E, Supplementary Figure 1**), despite different sampling approaches used for sequencing. Specifically, genomes from San Francisco County were derived from COVID-19 patients being tested at University of California, San Francisco (UCSF) hospitals and clinics; genomes from Alameda County were derived from community testing; genomes from Santa Clara County were derived from congregate facility, community, and acute care testing; and genomes from Los Angeles County were derived from coroner, community, and inpatient testing.

**Figure 1.**
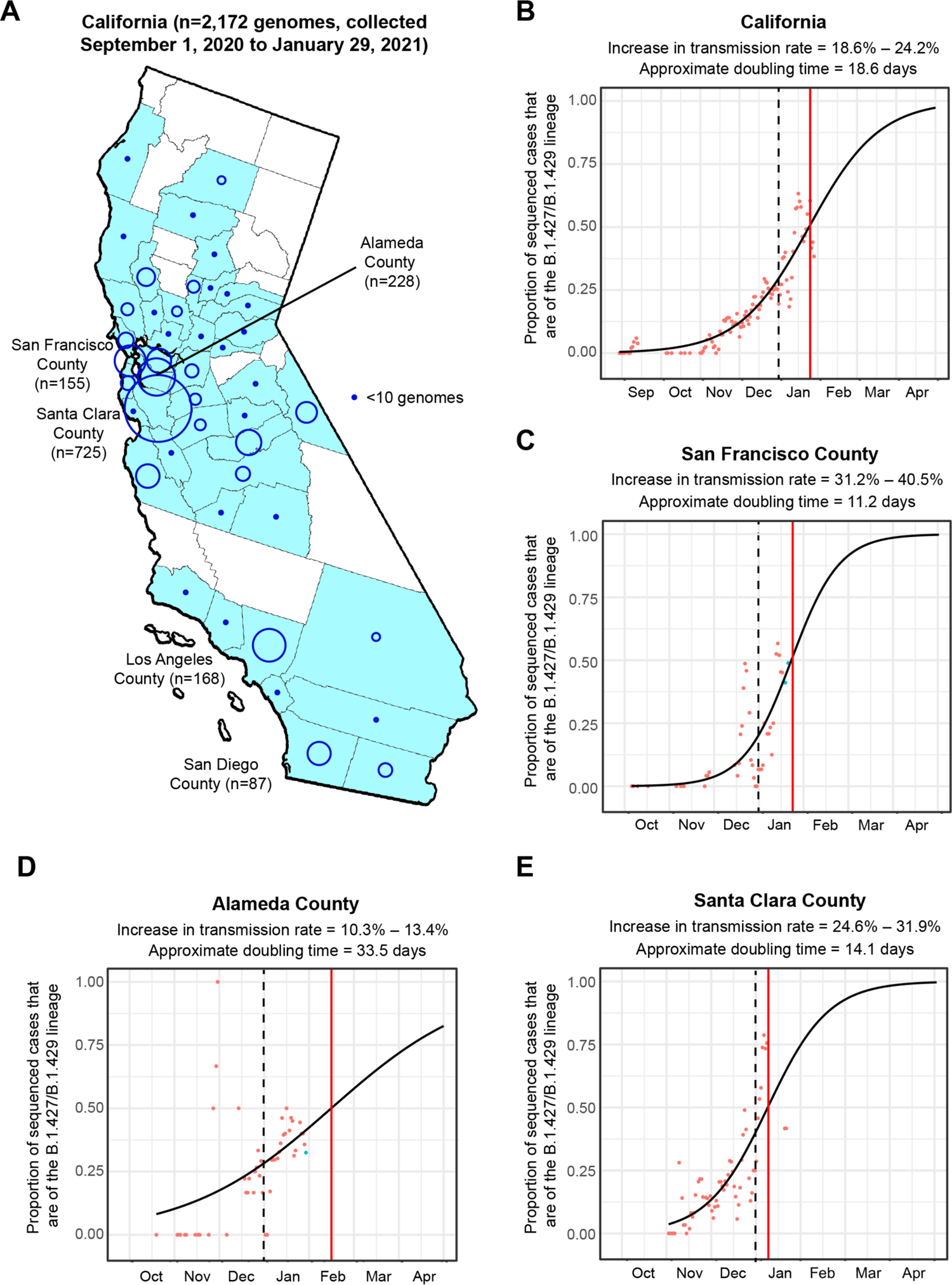
Increasing frequency of the B.1.427/B.1.429 variant in California from September 1. 2020 to January 29, 2021. **(A)** County-level representation of the 2,172 newly sequenced SARS-CoV-2 genomes in the current study. Counties from which at least 1 genome were sequenced are colored in sky blue. The size of the circle is proportionally to the number of genomes sequenced from each county, while points designate counties for fewer than 10 genomes were sequenced. Logistic growth curves fitting the 5-day rolling average of the estimated proportion of B.1.427/B.1.429 variant cases in **(B)** California, **(C)** San Francisco County, **(D)** Alameda County, and **(E)** Santa Clara County. The predicted time when the growth curve crosses 0.5 is indicated by a vertical red line. A vertical black dotted line denotes the transition from 2020 to 2021. The increase in transmission rate is defined as the change in the relative proportion of B.1.427/B.1.429 variant cases relative to circulating non-B.1.427/B.1.429 variant lineages as estimated from the logistic growth model (Volz et al., 2020; Washington et al., 2021).

### Phylogenetic and molecular dating analyses

Bayesian phylogenetic analysis of 1,166 genomes subsampled from a 2,519-genome dataset consisting of the 2,172 California genomes sequenced in this study and 347 representative global genomes (Bedford and Neher, 2020) identified two distinct lineages in clade 20C (Nextstrain designation) associated with the novel variant, B.1.427 and B.1.429 (**Figure 2B**). Both lineages share a triad of coding mutations in the spike protein (S13I, W152C, and L452R), one coding mutation in the orf1b protein (D1183Y), and an additional 2 non-coding mutations (**Figure 2A**). Four additional mutations, one of them a coding mutation orf1a:I4205V, were specific to B.1.429, while 3 additional non-coding mutations were specific to B.1.427. Using a previously reported algorithm to assess divergence time dating (Drummond et al., 2012), we estimated that the most recent common ancestor emerged on May 20, 2020 (95% highest posterior density [HPD] interval: April 29-June 9). The branches giving rise to the B.1.427 and B.1.429 lineages were predicted to have diverged on July 27 (95% HPD: June 6-September 8) and June 9 (95% HPD: May 23-June 23), respectively (**Figure 2C**).

**Figure 2.**
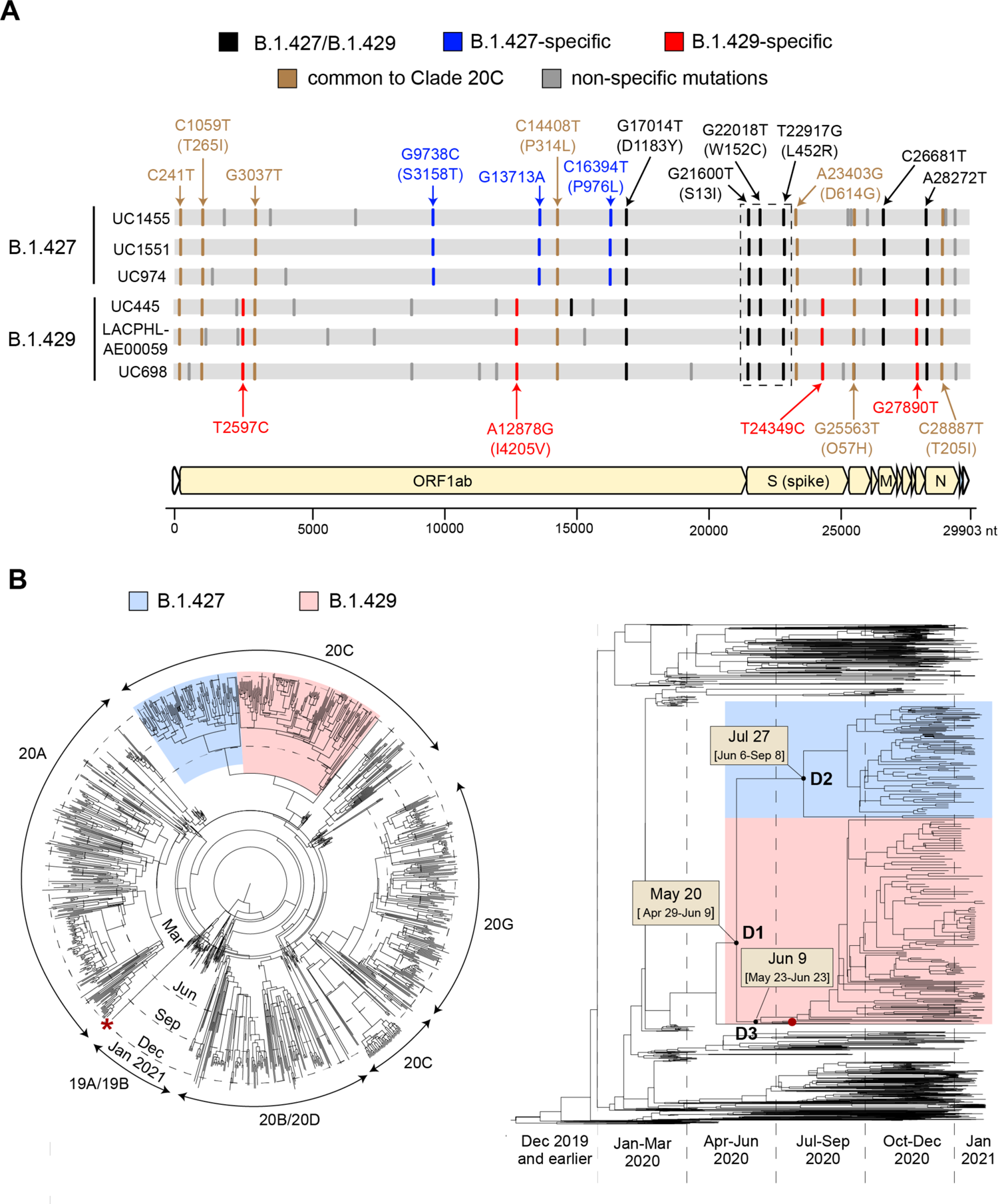
Genomic, phylogenetic, and molecular clock analyses of the B.1.427/B.1.429 variant in California. **(A)** A multiple sequence alignment of 6 representative B.1.427/B.1.429 genomes, 3 from the B.1.427 lineage and 3 from the B.1.429 lineage, using the prototypical Wuhan Hu-1 genome as a reference. Defining single nucleotide polymorphisms (SNPs) in the B.1.427 and B.1.429 lineages are compared to each other and to other SARS-CoV-2 viruses in Nextstrain clade 20C. The SNPs are color coded as follows: black SNPs are shared between the B.1.427 and B.1.429 lineages, blue SNPs are specific to B.1.427, red SNPs are specific to B.1.429, brown SNPs are shared among nearly all clade 20C viruses, and gray SNPs are specific to individual viruses. **(B)** Bayesian phylogenetic tree of 1,166 subsampled genomes constructed using molecular clock analysis from a complete dataset consisting of the 2,172 genomes recovered in the current study and 347 representative global genomes. The left panel shows a radial view of the tree, with marking of segments corresponding to the major clades. The right panel shows the divergence dates and associated 95% highest posterior density (HPD) distributions, or confidence intervals, for the B.1.427/B.1.429 variant (D1), B.1.427 lineage (D2), and B.1.429 lineage (D3), as estimated from TMRCA (time to most recent common ancestor) calculations. The B.1.427 lineage is colored in blue, and the B.1.429 lineage in red. The red asterisk denotes a UK B.1.1.7 variant genome. The red dot denotes the first reported genomic sequence of the B.1.429 variant from Los Angeles County from a sample collected July 13, 2020.

### Increased transmissibility and infectivity

Analysis of data from 2,126 (97.8%) of the 2,172 sequenced genomes in the current study revealed that the median PCR cycle threshold (Ct) value associated with B.1.427/B.1.429 variant infections was significantly lower (p=3.47×10^-6^) than that associated with non-variant viruses (**Figure 3C**). We estimated that in swab samples N/NP viral RNA is approximately 2-fold higher in B.1.427/B.1.429 than in non-variant viruses (Drew et al., 2020).

**Figure 3.**
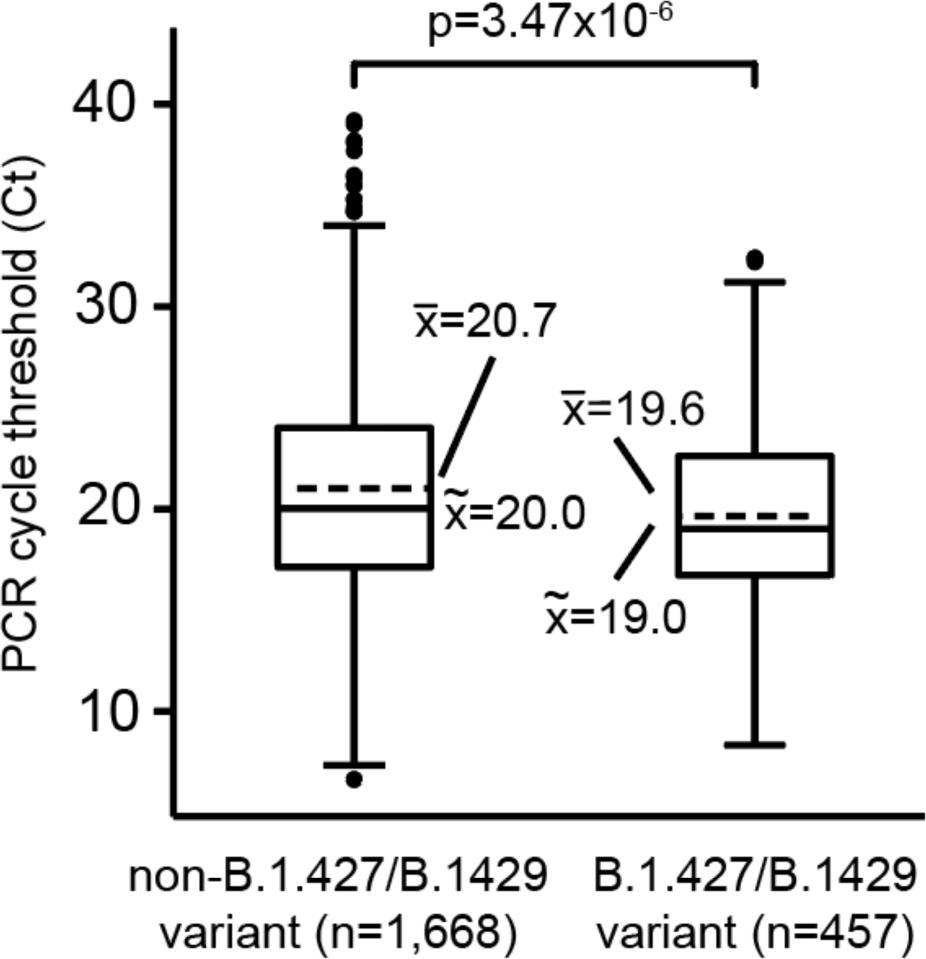
Higher viral loads in infections from the B.1.427/B.1.429 variant as compared to non-B.1.427/B.1.429 variant lineages. Box and whisker plots of available PCR cycle threshold (C_t_) values for B.1.427/B.1.429 variant (left) and non-variant (right) samples sequenced in the current study. Note that a C_t_ difference of 1 represents a 2-fold difference in the virus concentration (Drew et al., 2020). The dashed horizonal line in the box denotes the median value, the solid horizontal lines the mean value. The interquartile ranges are calculated with respect to the median value.

Analysis of the SARS-CoV-2 spike protein complexed to its human ACE2 receptor (Lan et al., 2020) revealed that the L452 residue does not directly contact the receptor. Instead, L452 together with F490 and L492 form a hydrophobic patch on the surface of the spike RBD (**Figure 4A**). To understand the effects of L452R RBD mutation on viral entry, pseudoviruses carrying D614G with L452R or W152C, or D614G alone were generated and used for infection of 293T cells stably expressing the ACE2 cell entry receptor and TMPRSS2 cofactor for SARS-CoV-2 (Hoffmann et al., 2020) and human airway lung organoids (HAO) stably expressing ACE2. We observed increased entry by pseudoviruses carrying the L452R mutation compared to D614G alone, with a 6.7 to 22.5-fold increase in 293T cells and a 5.8 to 14.7-fold increase in HAOs (**Figure 4B and 4C**). This increase in infection with L452R mutation is slightly lower than the increase observed with the N501Y mutation (11.4 to 30.9-fold increase in 293T cells and 23.5 to 37.8-fold increase in HAO relative to D614G alone), which has previously been reported to increase pseudovirus entry (Hu et al., 2021). Pseudoviruses carrying the W152C mutation demonstrated small increases in infection of 293T cells and HAO relative to the D614 control, although these increases were not as pronounced as those observed for the L452R and N501Y pseudoviruses.

**Figure 4.**
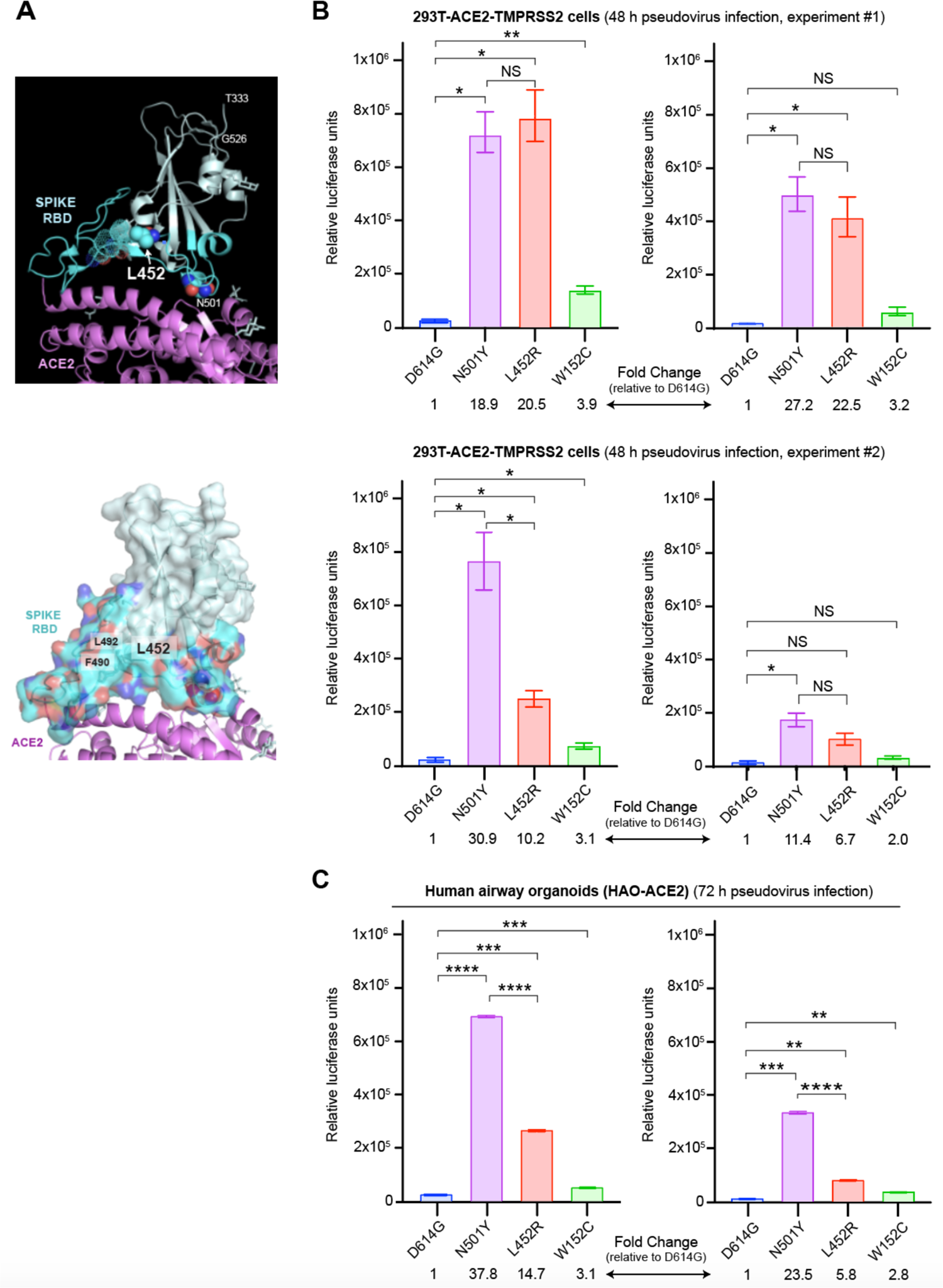
Increased infectivity of L452R-carrying pseudoviruses. **(A) *Upper panel:*** Ribbon diagram of the SARS-CoV-2 spike RBD in cyan bound to ACE2 receptor in magenta (PDB ID 6M0J). The receptor-binding motif of RBD is colored in dark cyan with L452 in solid spheres and F490 and L492 with dotted spheres. Sugars and Zn2+ are shown in grey. The position of N501 in direct contact with the ACE2 receptor is also shown for purposes of comparison. ***Lower panel:*** Surface representation of the spike RBD showing the hydrophobic patch outlined by L452, F490, and L492. (**B)** Levels of infection of SARS-CoV-2 spike pseudoviruses carrying D614G alone or D614G with N501Y, L452R, or W152C mutations in 293T cells stably expressing ACE2 and TMPRSS2. 293T cells were seeded in 96-well plates and infected with high (6 ng, **left**) or low (3 ng, **right**) concentrations of the indicated pseudoviruses for 48 h. Two biological replicates were assessed in two independent experiments, with 3 technical replicates per experiment. **(C)** Levels of infection in human lung airway organoids (HAO) stably expressing ACE2. HAO were seeded in 24-well plates and infected with high (4 ng, **left**) or low (2 ng, **right**) concentrations of the indicated pseudoviruses for 72 h. Pseudovirus cell entry was measured with a luciferase assay. The error bars represent the standard deviation of 3 technical replicates. Dunn’s multiple comparisons test was used to determine significance. Note that each of the N501Y, L452R, and W152G pseudoviruses also carries D614G. Abbreviations: NS, not significant.

### Reduced susceptibility to neutralizing antibodies from convalescent patients and vaccine recipients

To examine the effect of the L452R mutation on antibody binding, we performed neutralizing antibody assays. We cultured a B.1.429 lineage virus from a patient’s NP swab sample in Vero TMPRSS2 cells. We then performed plaque reduction neutralization tests (PRNT) using 21 plasma samples from convalescent patients and vaccine recipients to compare neutralization titers between the B.1.429 isolate and a control isolate USA-WA1/2020 (**Figure 5A**, **Supplementary Table 3**, and **Supplementary Figure 2**). Twelve samples were collected from individuals after receiving both doses of either the Pfizer BNT16b2 or Moderna mRNA-1273 vaccine, with samples collected 4-28 days after the second dose. Nine samples were convalescent plasma collected from patients clinically diagnosed with COVID-19 from August 20 to December 10, 2020, with samples collected 18 to 71 days after symptom onset. Measurable neutralizing antibody responses in the assay range were not observed for 1 convalescent patient and 1 vaccine recipient.

**Figure 5.**
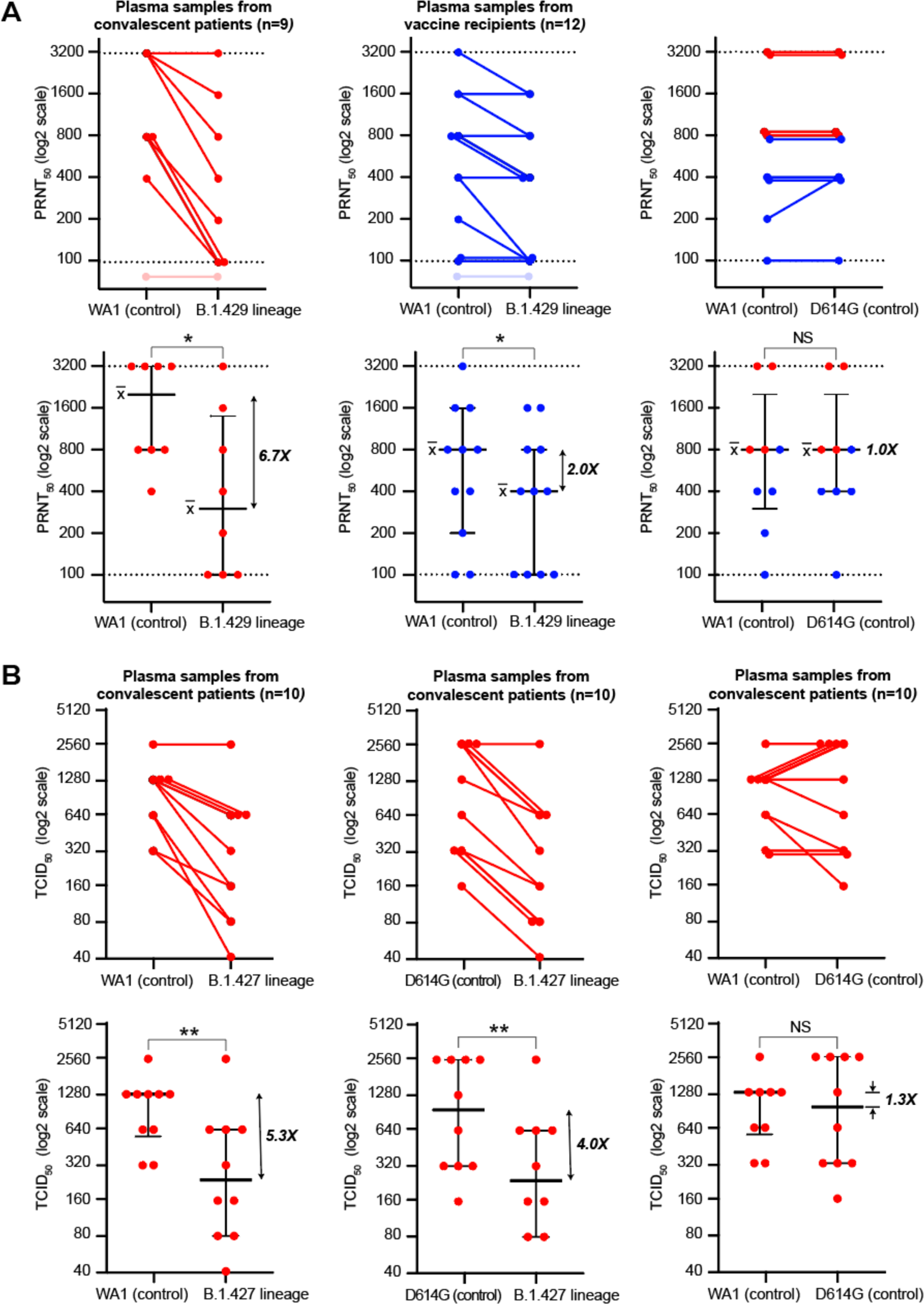
B.1.427/B.1.429 variant resistance to antibody neutralization in vitro. (A) Antibody neutralization titers from 9 convalescent patients and 12 vaccine recipients against cultured WA1 (control), D614G (control), and B.1.429 viral isolates were assessed using a PRNT assay. Lines connect the individual plasma samples tested pairwise for neutralization **(top row).** Only a subset of the plasma samples were tested with the WA1 and D614G head-to-head comparisons **(top row, right).** The dotted lines denote the upper and lower bounds for the PRNT assay (1:100 to 1:3200). Plasma samples that did not exhibit detectable neutralizing activity at titers above the lower threshold are shown as transparent. Individual PRNT_50_ measurements are plotted along with error bars denoting the median and standard deviation **(bottom row)**. **(B)** Antibody neutralization titers from 10 convalescent patients against cultured WA1 (control), D614G (control) and B.1.427 viral isolates were assessed by 50% CPE endpoint dilution. Lines connect the individual plasma samples tested pairwise for neutralization (**top row)**. Individual TCID_50_ measurements are plotted along with error bars denoting the median and standard deviation **(bottom row).** A Wilcoxon matched pairs signed rank test was used to determine significance. Abbreviations: NS, not significant; PRNT, plaque-reduction neutralization test; CPE, cytopathic effect; TCID, tissue culture infective dose.

We found that in comparison to USA-WA1/2020, 7 of 8 (88%) convalescent patients and 6 of 11 (55%) vaccine recipients, showed reduced PRNT_50_ titers to a B.1.429 lineage virus, with 6.7-fold (p=0.016) and 2-fold (p=0.031) median reductions, respectively (**Figure 5A**). There were no differences in neutralization between WA1 or D614G isolates by convalescent or post-vaccination plasma (**Figure 5A, right**).

Next, we independently evaluated neutralizing antibody responses against a cultured B.1.427 lineage virus. The TCID_50_, or median tissue culture infective dose at which 50% of cultures exhibit cytopathic effect (CPE), was determined for 10 different convalescent plasma samples collected from COVID-19 patients from June 19 to August 19, 2020, with samples collected 21 to 85 days after symptom onset. Nine of 10 (90%) convalescent patients showed reduced TCID_50_ titers to a B.1.427 lineage virus, with 5.3 (p=0.0039) and 4.0-fold (p=0.0039) median reductions for USA-WA1/2020) and D614G isolates, respectively.

## Discussion

As of early 2021, multiple SARS-CoV-2 variants have emerged in different regions of the world, each rapidly establishing itself as the predominant lineage within a few months after its initial detection (Chand et al., 2020; Sabino et al., 2021; Tegally et al., 2020). In the current study, we describe the spread of a novel B.1.427/B.1.429 variant in California carrying a characteristic triad of spike protein mutations (S13I, W152C, and L452R) that is predicted to have emerged in May 2020 and increased in frequency from 0% to >50% of sequenced cases from September 2020 to January 2021. Importantly, this variant was found to comprise 2 separate lineages, B.1.427 and B.1.429, with each lineage rising in parallel in California as well as in multiple other states (Gangavarapu et al., 2020). We also observed a moderate resistance to neutralization by antibodies elicited by prior infection (4.0 to 6.7-fold) or vaccination (2-fold). These findings indicate that the B.1.427/B.1.429 variant warrants close monitoring and further investigation regarding its potential to cause future surges in COVID-19 cases, accumulate further mutations, and/or decrease vaccine efficacy.

The results here highlight the urgent need for implementation of a robust genomic surveillance system in the US and globally to rapidly identify and monitor SARS-CoV-2 variants. Although our findings suggest that the B.1.427/B.1.429 variant emerged as early as May 2020, the first cases of B.1.427 and B.1.429 in the US were not identified by sequencing until September 28, 2020, and July 13, 2020, respectively. Sparse genomic sequencing of circulating viruses likely contributed to delayed identification of the B.1.427/B.1.429 variant. Furthermore, unlike in countries such as the UK (, 2020) and South Africa (Msomi et al., 2020), the US lacks an organized system for real-time analysis and reporting of variants that is tied to actionable public health responses. The first public disclosure of the existence of this variant, initiated by us in coordination with local and state public health agencies and the US CDC, did not occur until January 17, 2021 (CDPH, 2021), by which time the variant had already become the dominant lineage in several California counties and spread to multiple other states (Gangavarapu et al., 2020). Earlier identification and monitoring of the variant may have guided focused contact tracing efforts by public health to slow its spread, as well as enabled more timely investigation of its potential significance. Our identification of the B.1.427/B.1.429 variant was made possible by California COVIDNet, a collaborative sequencing network working to track transmission and evolution of SARS-CoV-2 in the state by viral whole-genome sequencing (CDPH, 2021).

The B.1.427/B.1.429 variant carries 4 new coding mutations, including 3 in the spike protein, that are not found in the 3 SARS-CoV-2 VOCs (B.1.1.7, B.1.351, and P.1) or in other major circulating lineages. The sudden appearance of several new mutations in a new variant is not unexpected. Indeed, the B.1.1.7 and B.1.351 variants each carry over 8 missense mutations in the spike protein (Rambaut et al., 2020b; Tegally et al., 2020). The evolutionary mechanism underlying the unusual genetic divergence of these emerging variants, with the accumulation of many mutations over a short time period, remains unexplained, but this divergence may potentially be due to accelerated viral quasispecies evolution in chronically infected patients (Avanzato et al., 2020; Choi et al., 2020; Kemp et al., 2021). Another possible explanation for the absence of genomes directly ancestral to B.1.427/B.1.429 is the aforementioned limited genomic sampling of SARS-CoV-2 in California and the US to date.

Prior studies have suggested that the L452R mutation may stabilize the interaction between the spike protein and its human ACE2 receptor and thereby increase infectivity (Chen et al., 2020; Teng et al., 2020). Our findings of enhanced infection of 293T cells and lung organoids by pseudoviruses carrying L452R confirm these early predictions. Notably, the L452 residue does not directly contact the ACE2 receptor, unlike the N501 residue that is mutated to Y501 in the highly transmissible B.1.1.7, B.1.351 and P.1 variants (**Figure 4A**). However, given that L452 is positioned in a hydrophobic patch of the spike RBD, it is plausible that the L452R mutation causes structural changes in the region that promote the interaction between the spike protein and its ACE2 receptor. Notably, our findings reveal that the infectivity of L452R pseudoviruses was higher than D614G, but slightly reduced compared to that of N501Y pseudoviruses in 293T cells and human airway lung organoids. Thus, whether the L452R-carrying B.1.427/B.1.429 will continue to remain the predominant circulating strain in California, or whether it will eventually be replaced by the N501Y-carrying B.1.1.7 variant (Washington et al., 2021) remains unclear.

The L452R mutation in the B.1.427/B.1.429 variant has been observed previously in rare, mostly singleton cases, first reported from Denmark on March 17, 2020, and also reported from multiple US states and the UK prior to September 1, 2020 (Gangavarapu et al., 2020). Given our findings of increased infectivity of L452R pseudoviruses, it is unclear why surges in L452R-carrying lineages have not occurred earlier. We speculate that although these lineages may have been more infective, transmission may not have reached a critical threshold locally or may have been influenced by other factors such as population density and/or public health interventions. An alternative (but not mutually exclusive) possibility is that the additional mutations in B.1.427/B.1.429, especially the W152C and S13I mutations in the spike protein, may contribute to increased infectivity of the variant relative to lineages carrying the L452R mutation alone. Indeed, in the current study we observed smaller but statistically significant increases in infection of 293T cell and lung organoids by pseudoviruses carrying W152C. Studies of pseudoviruses carrying the 3 spike mutations or the full complement of mutations in the B.1.427/B.1.429 variant are needed to address these hypotheses.

Our neutralization findings are consistent with a prior report showing decreased binding of L452R-carrying pseudoviruses by antibodies from previously infected COVID-19 patients and escape from neutralization in 3 of 4 convalescent plasma samples (Liu et al., 2020). We speculate that mutation of the L452 residue in a hydrophobic pocket may induce conformational changes in the RBD that impact neutralizing antibody binding. Of note, a >4-fold decrease in neutralizing antibody titers in convalescent plasma suggest that immune selection pressure from a previously exposed population may be partly driving the emergence of L452R variants. These data also raise questions regarding potential higher risk of re-infection and the therapeutic efficacy of monoclonal antibodies and convalescent plasma to treat COVID-19 disease from the B.1.427/B.1.429 variant.

Overall, the modest 2-fold decrease in neutralizing antibody titers in vaccine recipients to the B.1.429 variant is an indication of the robust neutralizing antibody responses elicited by mRNA vaccines in the face of variants under immune selection pressure. Indeed, a reduction in neutralization associated with the L452R mutation has been reported following vaccination, although the observed decrease in neutralizing antibody titers was similar at 2.9-fold (Garcia-Beltran et al., 2021). The use of a B.1.429 isolate in the present study, carrying the full complement of mutations that characterize the lineage, may account for some of the differences between the Garcia-Beltran et al. study and ours, and the contribution of epistatic mutations to neutralization phenotypes for SARS-CoV-2 variants merits further study. In addition, as neutralizing antibodies in natural infection have been shown to wane over time (Lau et al., 2021; Seow et al., 2020), longitudinal serologic studies are needed to determine whether these modest decreases will affect the long-term durability of vaccine-elicited immune responses to the B.1.427/B.1.429 variant. Of concern is also the possibility that B.1.427/B.1.429 lineages may accumulate additional mutations in the future that may further enhance the escape phenotype.

## Data Availability

Assembled SARS-CoV-2 genomes in this study were uploaded to GISAID (Elbe and Buckland-Merrett, 2017; Shu and McCauley, 2017) (accession numbers in Supplementary Table 1) and can be visualized in NextStrain. Viral genomes were also submitted to the National Center for Biotechnology Information (NCBI) GenBank database (accession numbers pending). Raw sequence data were submitted to the Sequence Read Archive (SRA) database (BioProject accession number PRJNA171119, Chiu laboratory at UCSF; BioProject accession number PRJNA639591, Wyman laboratory at UC Berkeley).

## Acknowledgments

We acknowledge the help from Delsy Martinez and Tyler Miyasaki at UCSF CAT core facility for genome sequencing efforts using NovaSeq. We would also like to acknowledge Maria Salas, Elizabeth Baylis and the entire COVIDNet team at the Viral and Rickettsial Disease Laboratory of the California Department of Public Health. We would also like to thank the Whelan lab at the Washington University School of Medicine for the Vero TMPRSS2 cell line. Author credits for specific GISAID contributions can be found on https://www.gisaid.org/. The findings and conclusions in this article are those of the author(s) and do not necessarily represent the views or opinions of the California Department of Public Health or the California Health and Human Services Agency.

## Author contributions

CYC, MO, and RA conceived and designed the study. CYC, XD, MAG-K, VS, CW, and GRK coordinated the sequencing efforts and laboratory studies. XD, MAG-K, MMK, VS, CW, AS-G, DRG, KRR, CSSM, BS, P-YC, US-G, TYT, JMH, CRS, PVL, YX, and MKM performed experiments. CYC, SF, and XD assembled and curated viral genomes. CYC performed the phylogenetic and molecular clock analyses. CYC, XD, MAG-K, VS, CW, KRR, ASG, NPR, JB, JT, JC, GRK, and CYC analyzed data. VS, CW, AS-G, ASG, NPR, KRR, JAS, and SM collected and sequenced SARS-CoV-2 samples from UCSF and throughout California. PH and NMG collected and sequenced samples from Los Angeles County. CA and DF collected and sequenced samples from Monterey County. FL, PAF, HS, and SKW collected and sequenced samples from Alameda County. CYC, XD, MAGK, VS, and CW wrote the manuscript. CYC, MAGK, GRK, and VS prepared the figures. CYC, XD, MAGK, VS, DAW, JKH, and CW edited the manuscript. All authors read the manuscript and agree to its contents.

## Resource Availability Lead Contact

Further information and requests for resources and reagents should be directed to and will be fulfilled by the Lead Contact, Charles Chiu.

## Materials Availability

This study did not generate any new reagents.

## Cell lines for virus culture

Vero E6 cells and Vero cells expressing human TMPRSS2 were used for SARS-CoV-2 viral culture. The culture was maintained in a humidified incubator at 37°C in 5% CO_2_ in the indicated media and passaged every 3-4 days.

## Methods

### Sample collection and diagnostic assay of SARS-CoV-2

Remnant nasal/nasopharyngeal (N/NP) swab samples in universal transport media (UTM) or viral transport media (VTM) (Copan Diagnostics, Murrieta, CA, USA) from RT-PCR positive COVID-19 patients were obtained from the University of California, San Francisco (UCSF) Clinical Microbiology Laboratory, the Innovative Genomics Institute (IGI) at University of California, Berkeley, California Department of Public Health, Santa Clara County and Los Angeles County for SARS-CoV-2 genome sequencing. A small fraction of swab samples (<1%) were obtained from the anterior nares. Clinical samples from state and county public health laboratories were collected and sequenced as part of routine public health surveillance activities. Clinical samples from the IGI were sequenced under a waiver from the UC Berkeley Office for the Protection of Human Subjects. Clinical samples from UCSF were collected for a biorepository and sequenced according to protocols approved by the UCSF Institutional Review Board (protocol number 10-01116, 11-05519).

Due to variation in results reported by different clinical testing platforms used at UCSF, the Taqpath™ Multiplex Real-time RT-PCR test, which includes nucleoprotein (N) gene, spike (S) gene, and orf1ab gene targets, was used to determine cycle threshold (Ct) values for PCR-positive samples. The Taqpath™ assay was also used for determining Ct values for PCR-positive samples from Alameda County that were sequenced by the University of California, Berkeley IGI and from the California Department of Public Health.

### Viral Genome Sequencing

NP swab samples were prepared using 100 uL of the primary sample in UTM or VTM mixed with 100uL DNA/RNA shield (Zymo Research, # R1100-250). The 1:1 sample mixture was then extracted using the Omega BioTek MagBind Viral DNA/RNA Kit (Omega Biotek, # M6246-03) on KingFisherTM Flex Purification System with a 96 deep-well head (ThermoFisher, 5400630). Extracted RNA was reverse transcribed to complementary DNA and tiling multiplexed amplicon PCR was performed using SARS-CoV-2 primers Version 3 according to a published protocol (Quick et al., 2017). Amplicons were ligated with adapters and incorporated with barcodes using NEBNext Ultra II DNA Library Prep Kit for Illumina (New England Biolabs, # E7645L). Libraries were barcoded using NEBNext Multiplex Oligos for Illumina (96 unique dual-index primer pairs) (New England Biolabs, # E6440L) and purified with AMPure XP (Beckman-Coulter, #. Amplicon libraries were then sequenced on either Illumina MiSeq or Novaseq 6000 as 2×150 paired-end reads (300 cycles).

### Genome Assembly and Variant Calling

Genome assembly of viral reads and variant calling were performed using an in-house developed bioinformatics pipeline as previously described (Deng et al., 2020). In short, Illumina raw paired-end reads were first screened for SARS-CoV-2 sequences using BLASTn (BLAST+ package 2.9.0) alignment against viral reference genome NC_045512, and then processed using the BBTools suite, v38.87 (Bushnell, 2021). Adapter sequences were trimmed and low-quality reads were removed using BBDuk, and then mapped to the NC_045512 reference genome using BBMap. Variants were called with CallVariants and a depth cutoff of 5 was used to generate the final assembly. A genome coverage breadth of >=70% was required for inclusion in the study.

Multiple sequence alignment of 6 B.1.427/B.1.429 genomes and the Wuhan Hu-1 prototypical genome (GISAID ID: EPI_ISL_402125, GenBank accession number MN908947) was performed using the MAFFT aligner v7.388 (Katoh and Standley, 2013) as implemented in Geneious v11.1.5 (Kearse et al., 2012).

### Phylogenetic Analysis

High-quality SARS-CoV-2 genomes (n=2,519, 2,172 generated in the current study and 347 used as representative global genomes) were downloaded from the Global Initiative on Sharing of All Influenza Data (GISAID) database and processed using the NextStrain bioinformatics pipeline Augur using IQTREE v1.6. Branch locations were estimated using a maximum-likelihood discrete traits model. The resulting tree was visualized in the NextStrain web application Auspice and in Geneious v11.1.5 (Kearse et al., 2012). Molecular clock analysis of SARS-CoV-2 for estimating the TMRCA (time to most recent common ancestor) and divergence dates for the B.1.426/B.1.427 variant was performed using the Markov chain Monte Carlo (MCMC) method implemented by Bayesian Evolutionary Analysis on Sampling Trees (BEAST) software v.2.63 (Drummond et al., 2012). Briefly, a HKY85 nucleotide substitution model was used, using a strict clock model and exponential population growth. All models were run using default priors except for the exponential growth rate (Laplace distribution) for which the scale was set to 100. The chain length was set to 10 million states with a 10% burn-in. Convergence was evaluated using Tracer v1.7.1 (Rambaut et al., 2018). The resulting maximum clade credibility (MCC) tree was generated using TreeAnnotator v.2.6.3 (Drummond et al., 2012) and visualized using FigTree v.1.4.4 (Rambaut, 2021).

## Cell culture

Cells were maintained in a humidified incubator at 37°C in 5% CO_2_ in the indicated media and passaged every 3-4 days. Vero E6 cells were cultured in MEM supplemented with 1x penicillin-streptomycin-glutamine (Gibco) and 10% fetal calf serum (FCS). Vero cells overexpressing human TMPRSS2 were a kind gift from the Whelan lab (Case et al., 2020) and were maintained in DMEM supplemented with 1x sodium pyruvate, 1x penicillin-streptomycin-glutamine and 10% FCS.

## SARS-CoV-2 isolation and passages

For the B.1.429 neutralization studies, a non-B.1.427/B.1.429 variant SARS-CoV-2/human/USA/CA-UCSF-0001C/2020 clinical isolate carrying the D614G spike mutation was isolated as previously described (Samuel et al., 2020) and passaged in A549-ACE22 expressing cells. For isolation of the B.1.429 lineage virus, 100 μL of a NP swab sample from a COVID-19 patient that was previously sequenced and identified as B.1.429 was mixed 1:1 with serum free DMEM (supplemented with 1x sodium pyruvate and 1x penicillin-streptomycin-glutamine), and two-fold serial dilutions were made of the sample over six wells of a 96-well plate. 100 μL of freshly trypsinized Vero TMPRSS2 cells resuspended in DMEM (supplemented with 1x sodium pyruvate, 2x penicillin-streptomycin-glutamine, 5 μg/mL amphotericin B and 10% FCS) was added to each well and mixed. The culture was incubated at 37°C in 5% CO_2_ for 4-6 days and cytopathic effect (CPE) on cells was evaluated daily using a light microscope. The contents of wells positive for CPE were collected and stored at −80°C as a passage 0 stock (P0). P1 stocks were made following infection of four near confluent wells of a 24-well plates with Vero TMPRSS2 using the P0 stock. Supernatants were harvested 48 hours later after centrifugation at 800g for 7 minutes. P2 stocks were similarly made after infection of a near confluent T25 plate seeded with Vero E6 cells. All steps for isolation of the B.1.429 lineage virus were done in a Biosafety Level 3 lab using protocols approved by the Institutional Biosafety Committee at UCSF.

For the B.1.427 neutralization studies, Vero-81 cells were cultured with MEM supplemented with 1x penicillin-streptomycin (Gibco) and glutamine (Gibco) and 5% FCS (Hyclone). For isolation of B.1.427 and non-B.1.427/B.1.429 variant D614G viruses, 100 μL each from NP swab samples from COVID-19 patients identified as being infected by the B.1.427 or non-B.1.427/B.1.429 variant D614G lineage was diluted 1:5 in PBS supplemented with 0.75% bovine serum albumin (BSA-PBS) and added to confluent Vero-81 cells in a T25 flask. After adsorption for 1 h, additional media was then added, and the flask was incubated at 37°C with 5% CO_2_ for 3-4 days with daily monitoring for CPE. The contents were collected, clarified by centrifugation and stored at −80C as passage 0 stock. P1 stock was made by inoculation of Vero-81 confluent T150 flasks with 1:10 diluted p0 stock and similarly monitored and harvested to approximately 50% confluency. All steps for isolation of the B.1.427 lineage virus were done in a Biosafety Level 3 lab at the Viral and Rickettisial Disease Laboratory (VRDL) at the California Department of Public Health (CDPH).

For both the B.1.429 and B.1.427 neutralization studies, the SARS-CoV-2 USA-WA1/2020 strain (BEI resources) was passaged in Vero E6 cells (ATCC CRL-1586) or Vero-81 cells and used as a control. All stocks were resequenced and the consensus assembled viral genomes were identical to the genomes derived from the primary NP samples and carried all of the expected mutations.

## Plaque reduction neutralization tests using a B.1.429 lineage virus

Conventional PRNT assays were done using P2 stocks of B.1.429 lineage viruses and the USA-WA1/2020 isolate passaged on Vero E6 cells. Patient plasma was heat inactivated at 56°C for 30 minutes, clarified by centrifugation at 10,000 relative centrifugal force (rcf) for 5 minutes and aliquoted to minimize freeze thaw cycles. Serial 2-fold dilutions were made of plasma in PBS supplemented with 0.75% bovine serum albumin (BSA). Plasma dilutions were mixed with ∼100 plaque forming units (pfu) of viral isolates in serum free MEM in a 1:1 ratio and incubated for 1 hr at 37°C. Final plasma dilutions in plasma-virus mixtures ranged from 1:100 to 1:3200. 250 μL of plasma-virus mixtures were inoculated on a confluent monolayer of Vero E6 cells in 6-well plates, rocked and incubated for 1 h in a humidified incubator at 37°C in 5% CO_2_. After incubation, 3 mL of a mixture of MEM containing a final concentration of 2% FCS, 1x penicillin-streptomycin-glutamine and 1% melted agarose, maintained at ∼56°C, was added to the wells. After 72 h of culture as above, the wells were fixed with 4% paraformaldehyde for 2 h, agarose plugs were removed, and wells were stained with 0.1% crystal violet solution. Plaques were counted and the PRNT_50_ values were defined as the serum dilution at which 50% or more of plaques were neutralized. Assays were done in duplicate, and a positive control and negative control were included using plasma with known neutralizing activity (diluted 1:50) and from a SARS-CoV-2 unexposed individual (1:20 dilution), respectively. All steps were done in a Biosafety Level 3 lab using protocols approved by the Institutional Biosafety Committee at UCSF.

## CPE endpoint neutralization assays using a B.1.427 lineage virus

CPE endpoint neutralization assays were done following the limiting dilution model (Wang et al., 2005) and using P1 stocks of B.1.427, D614G, and USA-WA1/2020 lineages. Convalescent patient plasma was diluted 1:10 and heat inactivated at 56°C for 30 min. Serial 2-fold dilutions of plasma were made in BSA-PBS. Plasma dilutions were mixed with 100 TCID_50_ of each virus diluted in BSA-PBS at a 1:1 ratio (220 μL plasma dilution and 220 μL virus input) and incubated for 1 hour at 37C. Final plasma dilutions in plasma-virus mixture ranged from 1:20 to 1:2560. 100 μL of the plasma-virus mixtures were inoculated on confluent monolayer of Vero-81 cells in 96-well plates in quadruplicate and incubated at 37°C with 5% CO_2_ incubator. After incubation 150 μL of MEM containing 5% FCS was added to the wells and plates were incubated at 37°C with 5% CO_2_ until consistent CPE was seen in virus control (no neutralizing plasma added) wells. Positive and negative controls were included as well as cell control wells and a viral back titration to verify TCID_50_ viral input. Individual wells were scored for CPE as having a binary outcome of ‘infection” or ‘no infection’. The TCID_50_ was calculated as the dose that produced cytopathic effect in >50% of the inoculated wells. All steps were done in a Biosafety Level 3 lab using approved protocols.

## SARS-CoV-2 receptor binding domain mutagenesis and pseudovirus infection assay

SARS-CoV-2 spike mutants (D614G, D614G+W152C, D614G+L452R, and D614G+N501Y) were cloned using standard site-directed mutagenesis and PCR. Pseudoviruses typed with these spike mutants were generated as previously described with modifications (Crawford et al., 2020). Briefly, 293T cells were transfected with plasmid DNA (per 6-well plate: 340 ng of spike mutants, 1μg CMV-Gag-Pol (pCMV-dΔR8.91), 125 ng pAdvantage (Promega), 1 μg Luciferase reporter) for 48 h. Supernatant containing pseudovirus particles was collected, filtered (0.45μm), and stored in aliquots at −80°C. Pseudoviruses were quantified with a p24 assay (Takara #632200), and normalized based on titer prior to infection for entry assays. Human airway organoids (HAO) stably expressing ACE2 (HAO-ACE2) or 293T cells stably expressing ACE2 and TMPRSS2 (293T-ACE2-TMPRSS2) were infected with an equivalent amount of the indicated pseudoviruses in the presence of 5-10 ug/ml of polybrene for 72h. Pseudovirus entry was assayed using a luciferase assay (Promega #E1501) and luminescence was measured in a plate reader (TECAN, Infinite 200 Pro M Plex). Two independent experiments were run for the 293T pseudovirus assays (2 biological replicates), with 3 technical replicates run per experiment. The HAO pseudovirus assays were run as a single experiment with 3 technical replicates.

## Statistical analyses

The proportion of B.1.427/B.1.429 was estimated by dividing the number of B.1.427/B.1.429 variant cases by the total number of samples sequenced at a given location and collection date. A logistic growth curve fitting to the data points was generated using a non-linear least squares approach, as implemented by the nls() function in R(version 4.0.3). We estimated the increase in relative transmission rate of the B.1.427/B.1.429 variant by multiplying the logistic growth rate, defined as the change in the proportion of B.1.427/B.1.429 cases per day, by the serial interval (5 days for SARS-CoV2 (Rai et al., 2021)), as previously described (Volz et al., 2020; Washington et al., 2021). Similar to the analyses in Washington, et al., the doubling time was approximated using the formula: log (2) / logistic growth rate.

Welch’s t-test, as implemented in R (version 4.0.3) using the rstatix_0.7.0 package, was used to compare the N gene Ct values between B.1.427/B.1.429 variant and non-B.1.427/B.1.429 groups. For the in vitro pseudovirus infectivity studies, a one-way ANOVA test was used to determine significance. For the PRNT studies, a Wilcoxon matched pairs signed rank test was used to determine significance.

## Supplementary Figures and Tables

**Supplementary Figure 1.**
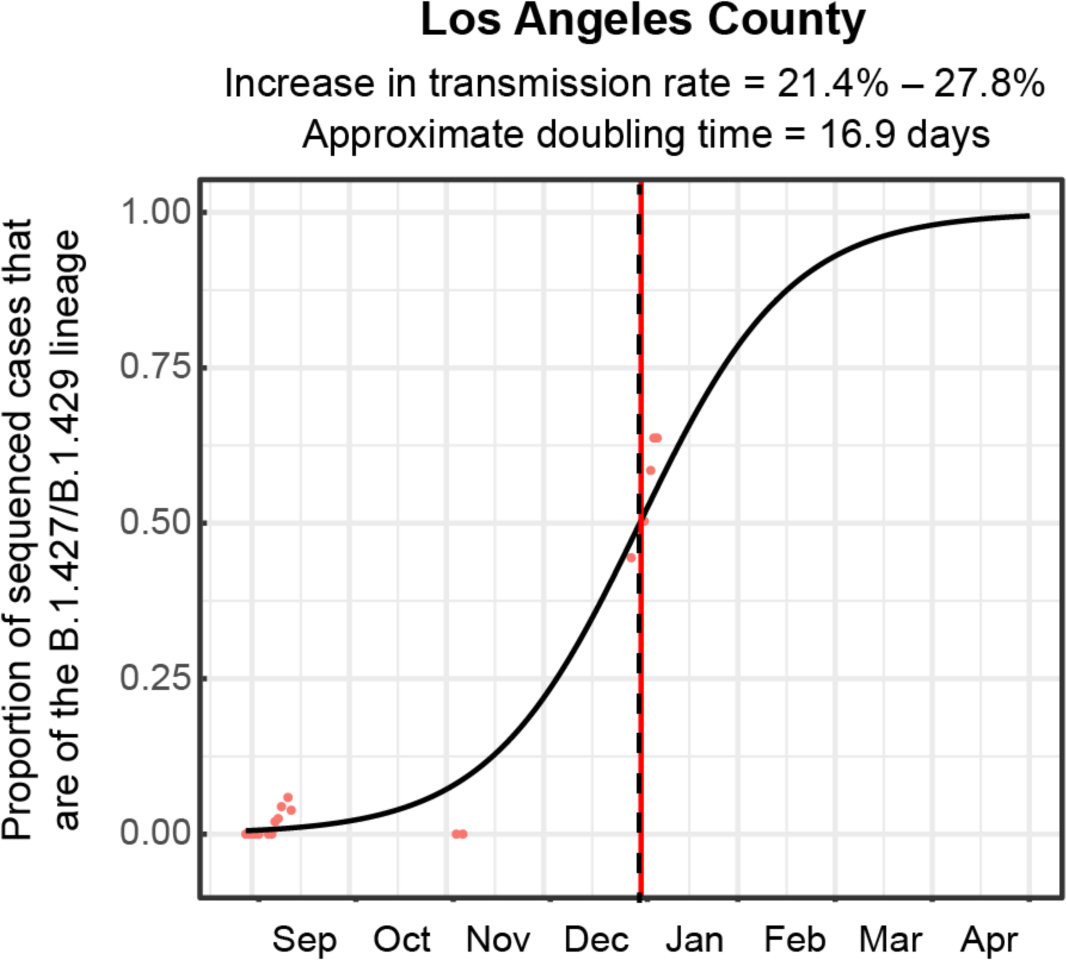
Increasing frequency of the B.1.427/B.1.429 variant in Los Angeles County from September 1. 2020 to January 29, 2021. Logistic growth curves fitting the 5-day rolling average of the estimated proportion of B.1.427/B.1.429 variant cases in Los Angeles County. A vertical black dotted line is used to denote the transition from 2020 to 2021.

**Supplementary Figure 2.**
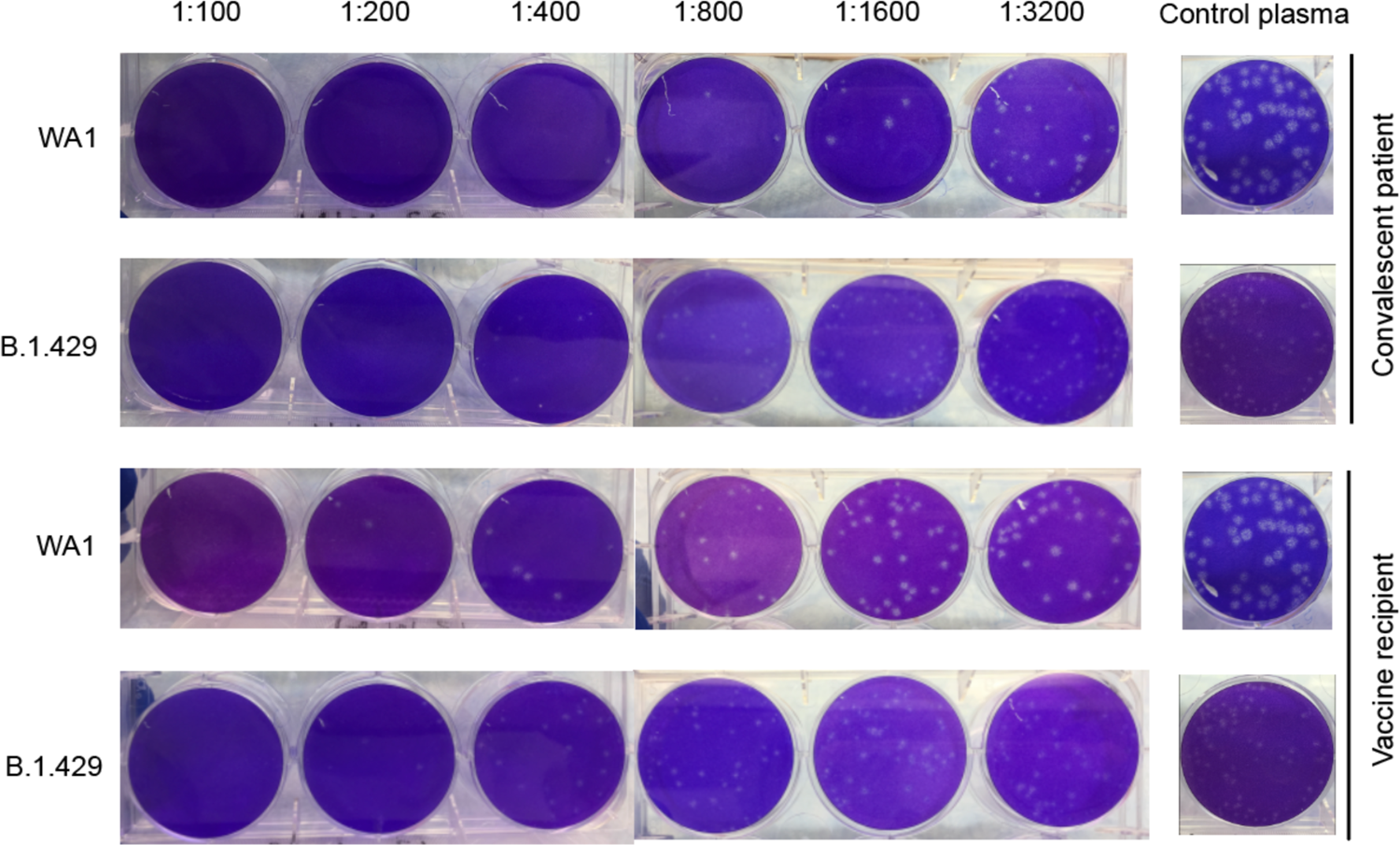
Differential neutralization of WA1 and B.1.429 viruses as measured by plaque-reduction neutralization tests. Representative 6-well plates arranged in one line showing viral plaques formed after co-culture with plasma samples from a convalescent patient and vaccine recipient. The same negative control well image is shown in line with the respective viral strain for both vaccine and convalescent samples. The plaques from B.1.429 lineage virus are observed to be small and lighter than those from control WA1 virus. The larger plaques for WA1 are likely due to adaptation in Vero E6 cells; these adaptation mutations have been reported not to impact neutralization responses (Klimstra et al., 2020).

**Supplementary Table 1. Metadata for the 2,172 genomes analyzed in this study.**

“SupplementaryTable1.xlsx”

**Supplementary Table 2.**
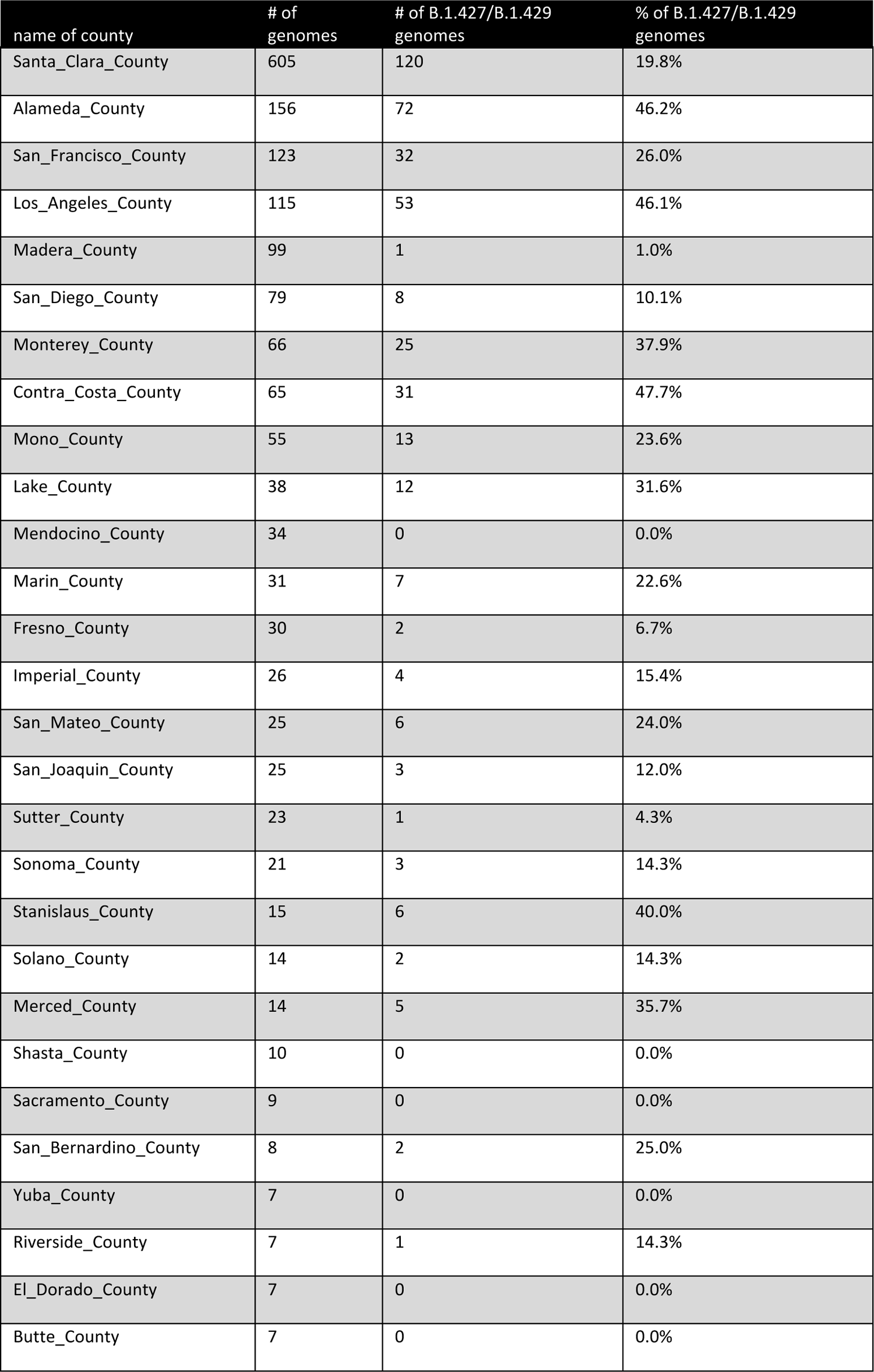
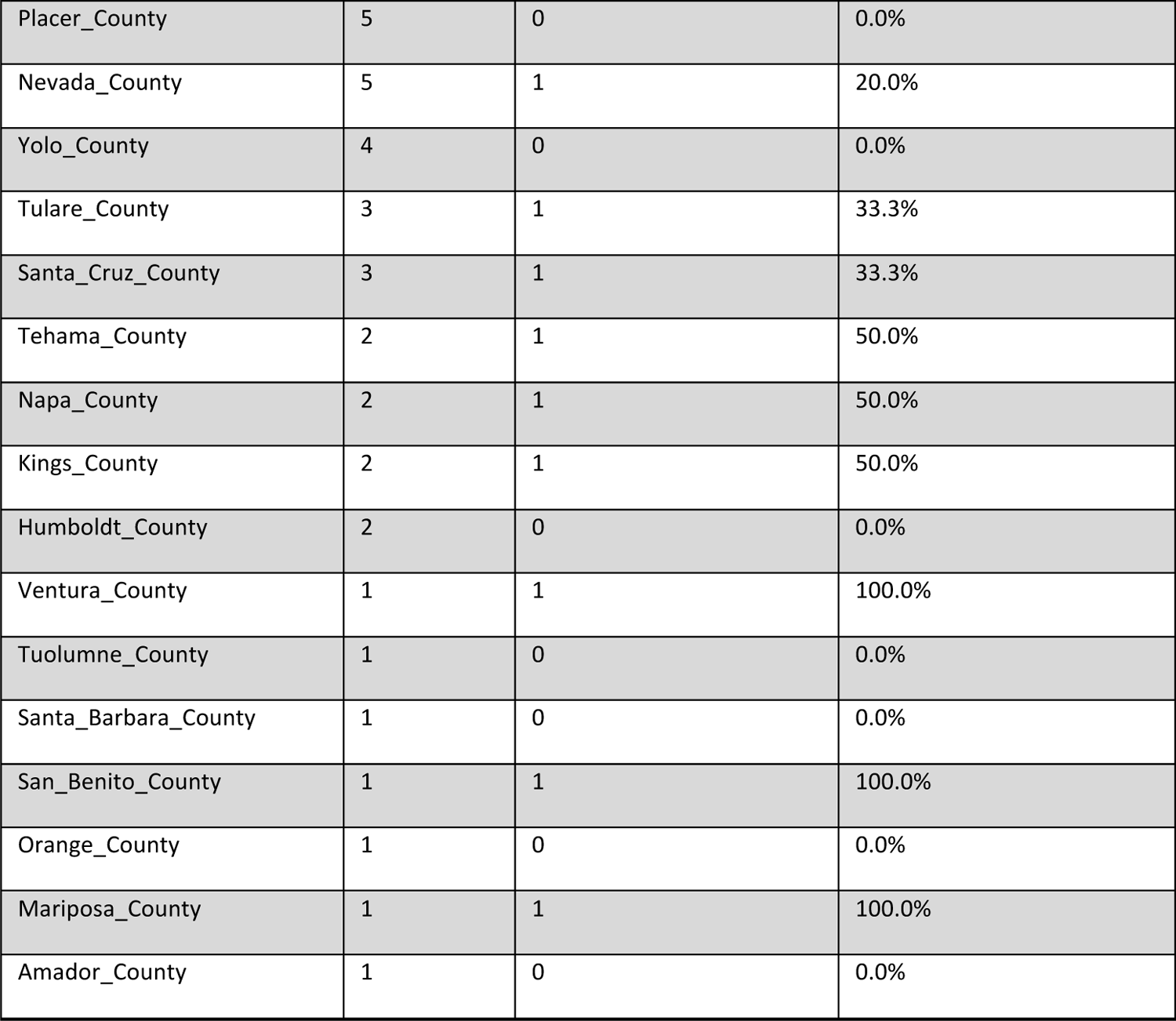
List of California counties and the B.1.427/B.1.429 genomes sequenced from each county

**Supplementary Table 3.**
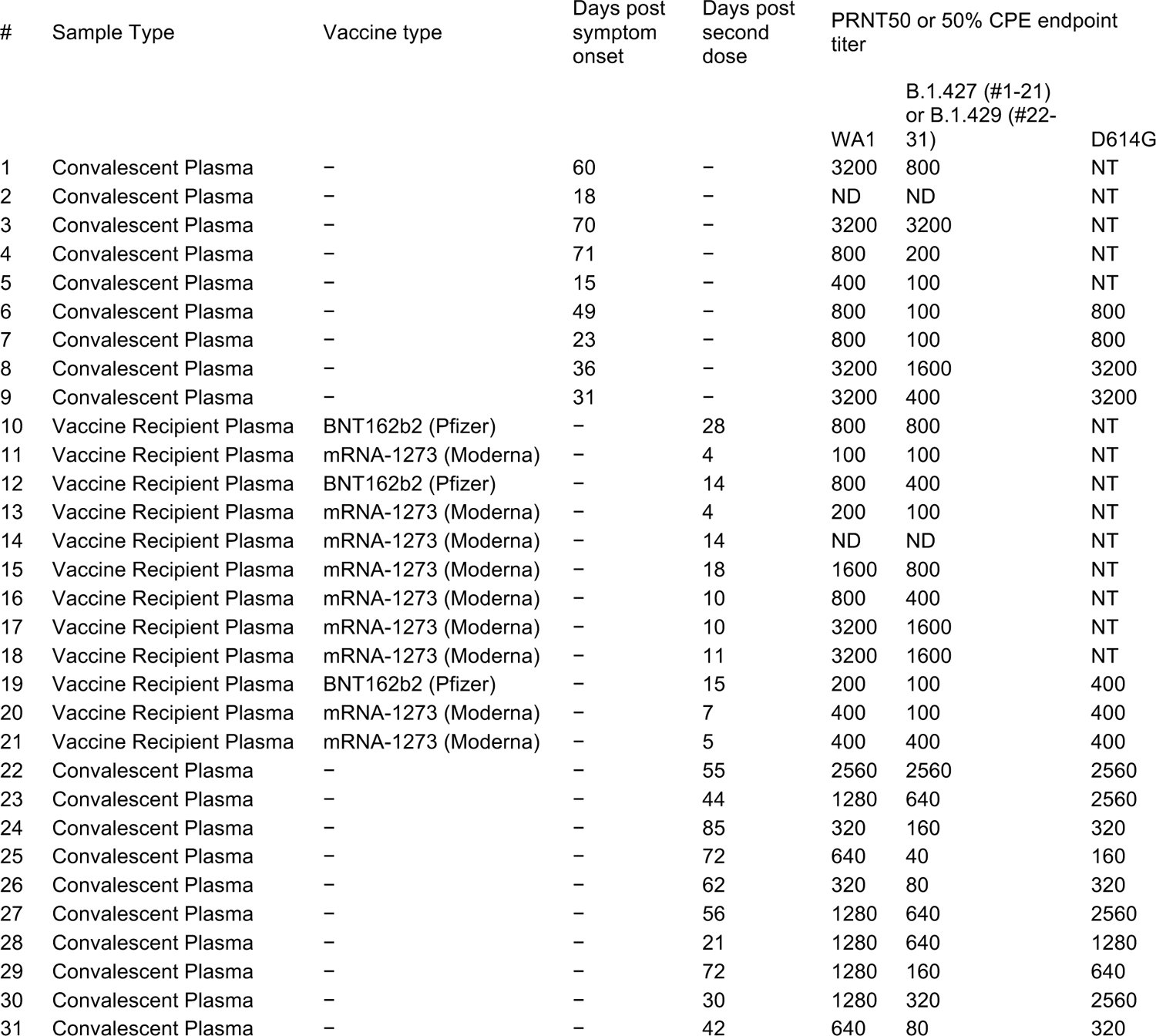
Plasma samples from convalescent COVID-19 patients and SARS-CoV-2 vaccine recipients used for evaluating neutralizing activity against B.1.427 and B.1.429 lineage viruses

